# Robust spike antibody responses and increased reactogenicity in seropositive individuals after a single dose of SARS-CoV-2 mRNA vaccine

**DOI:** 10.1101/2021.01.29.21250653

**Authors:** Florian Krammer, Komal Srivastava, the PARIS team, Viviana Simon

## Abstract

An important question is arising as COVID-19 vaccines are getting rolled out: Should individuals who already had a SARS-CoV-2 infection receive one or two shots of the currently authorized mRNA vaccines. In this short report, we show that the antibody response to the first vaccine dose in individuals with pre-existing immunity is equal to or even exceeds the titers found in naïve individuals after the second dose. We also show that the reactogenicity is significantly higher in individuals who have been infected with SARS-CoV-2 in the past. Changing the policy to give these individuals only one dose of vaccine would not negatively impact on their antibody titers, spare them from unnecessary pain and free up many urgently needed vaccine doses.

## Manuscript

Two SARS-CoV-2 spike mRNA vaccines received emergency use authorization by the FDA in December 2020 (BNT162b2/Pfizer; mRNA-1273/Moderna).^1^ Both Phase 3 trials reported high efficacy in preventing symptomatic SARS-CoV-2 infections after two doses of the vaccine administered three to four weeks apart (BNT162b2: 21 days; mRNA-1273: 28 days) in participants without previous COVID-19.^2,3^ For individuals with pre-existing immunity to SARS-CoV-2 the first vaccine dose likely immunologically resembles the booster dose in naïve individuals. Anecdotally, individuals with pre-existing immunity also experience more severe reactogenicity after the first doses compared to naïve individuals. This begs the question if individuals with pre-existing immunity should even receive a second dose of vaccine.

Here we describe the antibody responses in 109 individuals with and without documented pre-existing SARS-CoV-2 immunity (seronegative: 68, seropositive: 41) who received their first vaccine dose in 2020. Repeated sampling after the first dose indicates that the majority of seronegative individuals mount variable and relatively low SARS-CoV-2 IgG responses within 9-12 days after vaccination (median AUC pre-vaccination: 1 [N=68]; 9-12 days: 439 [N=13]; 13-16 days: 1037 [N=15], 17-20 days: 1,037 [N=15], 21-24 days: 1,075 [N=11], and post 2^nd^ dose 1,399 [N= 21]; Fig. 1A). In contrast, individuals with pre-existing SARS-CoV-2 immune responses (as evidenced by SARS-CoV-2 antibodies) rapidly develop uniform, high antibody titers within days of vaccination (median AUC pre vaccination: 91 [N=41]; 5-8 days: 14,208 [N=15], 9-12 days: 20,783 [N=8]; 13-16 days: 25,927 [N=20], 17-20 days: 12,661 [N=5], 21-24 days: 16,263 [N=4] and post 2^nd^ dose: 22,509 [N=7], Fig. 1A). The antibody titers of vaccinees with pre-existing immunity are not only 10-20 times higher than those of naïve vaccines at the same time points (p <0.0001, two tailed Mann Whitney test), but also exceed the median antibody titers measured in naïve individuals after the second vaccine dose by more than 10-fold. Ongoing follow-up studies will show whether these early differences in immune responses are maintained over time.

**Fig 1:**
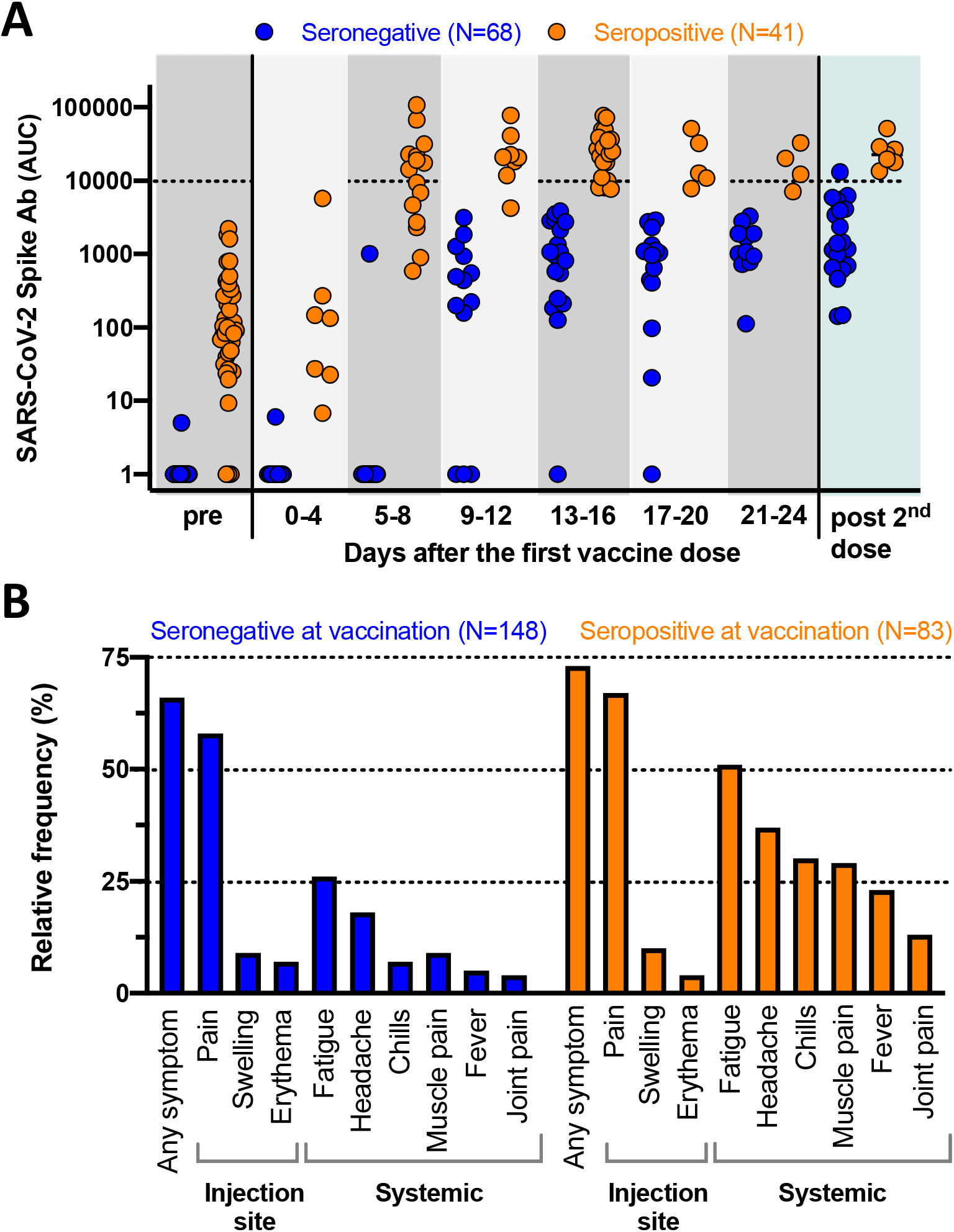
Immunogenicity and reactogenicity of SARS-CoV-2 RNA vaccines. A: Quantitative SARS-CoV-2 spike antibody titers (ELISA, expressed as area under the curve, AUC) for 109 individuals. “Pre” represents the antibody response prior to vaccination while “post 2^nd^ dose” indicates the immune responses mounted after the second vaccine dose. Note that some of the individuals with pre-existing immunity had antibody titers below detection (AUC of 1) at the time point prior to vaccination. B: Vaccine associated side effects experienced after the first dose (N= 231 individuals). The local side effects occur with comparable frequency while the systemic symptoms are significantly more common in the individuals with pre-existing immunity.

In addition, we compared frequency of local, injection side-related as well as systemic reactions after the first dose of vaccination in 231 individuals (148 seronegative and 83 seropositive; Fig. 1B). Overall both vaccines are well tolerated without any side effects requiring additional medical attention. 159/231 of the participants completing the survey after the first dose experienced any kind of side effect (66% seronegative and 73% seropositive). Most common were localized injection site symptoms (e.g., pain, swelling and erythema), which occurred with equal frequency independent of the serostatus at the time of vaccination and resolved spontaneously within days of vaccination. Vaccine recipients with pre-existing immunity experience systemic side effects with a significantly higher frequency than antibody naïve vaccines (e.g., fatigue, headache, chills, fever, muscle or join pains, in order of decreasing frequency, P < 0.001 for all listed symptoms, Fisher’s exact test, two-sided). Most of the participants for whom antibody results are presented above also completed the vaccine side-effect survey.

These findings suggest that a single dose of mRNA vaccine elicits very rapid immune responses in seropositive individuals with post-vaccine antibody titers that are comparable to or exceed titers found in naïve individuals who received two vaccinations. We also noted that vaccine reactogenicity after the first dose is substantially more pronounced in individuals with pre-existing immunity akin to side-effects reported for the second dose in the phase III vaccine trials^2,3^. These observations are in line with the first vaccine dose serving as boost in naturally infected individuals providing a rationale for updating vaccine recommendations to considering a single vaccine dose to be sufficient to reach immunity. Using quantitative serological assays that measure antibodies to the spike protein could be used to screen individuals prior to vaccination if the infection history is unknown.^4,5^ Such policies would allow not only expanding limited vaccine supply but also limit the reactogenicity experienced by COVID-19 survivors.

## Data Availability

The data will be shared upon request

## Acknowledgment

We thank the study participants for their generosity and continued support of COVID19 research.

## Ethics statement

The study protocols for the collection of clinical specimens from individuals with and without SARS-CoV-2 infection by the Personalized Virology Initiative were reviewed and approved by the Mount Sinai Hospital Institutional Review Board (IRB-16-00791; IRB-20-03374). All participants provided informed consent prior to collection of specimen and clinical information. All specimens were coded prior to processing.

## Conflict of interest statement

The Icahn School of Medicine at Mount Sinai has filed patent applications relating to SARS-CoV-2 serological assays and NDV-based SARS-CoV-2 vaccines which list Florian Krammer as co-inventor.

Daniel Stadlbauer and Viviana Simon are also listed on the serological assay patent application as co-inventors. Mount Sinai has spun out a company, Kantaro, to market serological tests for SARS-CoV-2. Florian Krammer has consulted for Merck and Pfizer (before 2020), and is currently consulting for Seqirus and Avimex. The Krammer laboratory is also collaborating with Pfizer on animal models of SARS-CoV-2.

## Funding statement

This work was partially funded by the NIAID Collaborative Influenza Vaccine Innovation Centers (CIVIC) contract 75N93019C00051, NIAID Center of Excellence for Influenza Research and Surveillance (CEIRS, contract # HHSN272201400008C), by the generous support of the JPB Foundation and the Open Philanthropy Project (research grant 2020-215611 (5384); and by anonymous donors.

## Notes

### Author Declarations

The study protocols for the collection of clinical specimens from individuals with and without SARS-CoV-2 infection by the Personalized Virology Initiative were reviewed and approved by the Mount Sinai Hospital Institutional Review Board (IRB-16-00791; IRB-20-03374). All participants provided informed consent prior to the collection of specimens and clinical information. All specimens were coded prior to processing.

